# Epigenetic and transcriptomic reprogramming in monocytes of severe COVID-19 patients reflects alterations in myeloid differentiation and the influence of inflammatory cytokines

**DOI:** 10.1101/2022.10.24.22281485

**Authors:** Gerard Godoy-Tena, Anis Barmada, Octavio Morante-Palacios, Carlos de la Calle-Fabregat, Ricardo Martins-Ferreira, Anna G. Ferreté-Bonastre, Laura Ciudad, Adolfo Ruiz-Sanmartín, Mónica Martínez-Gallo, Ricard Ferrer, Juan Carlos Ruiz-Rodriguez, Javier Rodríguez-Ubreva, Roser Vento-Tormo, Esteban Ballestar

## Abstract

COVID-19 manifests with a wide spectrum of clinical phenotypes, ranging from asymptomatic and mild to severe and critical. Severe and critical COVID-19 patients are characterized by marked changes in the myeloid compartment, especially monocytes. However, little is known about the epigenetic alterations that occur in these cells during hyperinflammatory responses in severe COVID-19 patients. In this study, we obtained the DNA methylome and transcriptome of peripheral blood monocytes from severe COVID-19 patients. DNA samples extracted from CD14+CD15-monocytes of 48 severe COVID-19 patients and 11 healthy controls were hybridized on MethylationEPIC BeadChip arrays. In parallel, single-cell transcriptomics of 10 severe COVID-19 patients were generated. CellPhoneDB was used to infer changes in the crosstalk between monocytes and other immune cell types. We observed DNA methylation changes in CpG sites associated with interferon-related genes and genes associated with antigen presentation, concordant with gene expression changes. These changes significantly overlapped with those occurring in bacterial sepsis, although specific DNA methylation alterations in genes specific to viral infection were also identified. We also found these alterations to comprise some of the DNA methylation changes occurring during myeloid differentiation and under the influence of inflammatory cytokines. A progression of DNA methylation alterations in relation to the Sequential Organ Failure Assessment (SOFA) score was found to be related to interferon-related genes and T-helper 1 cell cytokine production. CellPhoneDB analysis of the single-cell transcriptomes of other immune cell types suggested the existence of altered crosstalk between monocytes and other cell types like NK cells and regulatory T cells. Our findings show the occurrence of an epigenetic and transcriptional reprogramming of peripheral blood monocytes, which could be associated with the release of aberrant immature monocytes, increased systemic levels of pro-inflammatory cytokines, and changes in immune cell crosstalk in these patients.

## Background

Severe acute respiratory syndrome coronavirus 2 (SARS-CoV-2) causes the well-known Coronavirus disease 2019 (COVID-19), which has become a major global health burden. SARS-CoV-2 infection occurs through the nasopharyngeal mucosa[1]. Subsequent immune responses occur at the local mucosa and at a systemic level. An effective response to SARS-CoV-2 infection requires coordination between the innate and adaptive immune systems, including granulocytes, monocytes, macrophages, and T and B cells[2,3]. The range of immune responses to SARS-CoV-2 infection is diverse, from asymptomatic or mild upper-respiratory illness to severe viral pneumonia, acute respiratory distress syndrome and death[4]. The most severe forms of COVID-19 are caused by dysregulation of immune homeostasis, which leads to hyperinflammation in the lungs[5]. This has been shown to be more pronounced in the elderly and in individuals with pre-existing comorbidities[6,7]. Nevertheless, despite the numerous studies performed in the field, the impact of exacerbated immune responses associated with severe COVID-19 at the systemic level remains unclear.

Various studies have demonstrated that peripheral pathogenic T cells and inflammatory monocytes can induce a cytokine storm in severe COVID-19 patients[8]. This takes the form of excessive production of inflammatory mediators, specifically, interleukin (IL)-6, IL-1β, granulocyte-macrophage colony-stimulating factor (GM-CSF), tumor necrosis factor-alpha (TNFα) and interferon-gamma (IFNγ)[8–11]. IFN is essential for inducing the innate immune response during viral infection through different interferon regulatory factors (IRFs)[12]. Further, in COVID-19 patients, type I IFN deficiency appears to be a hallmark of severe cases[13–19] in association with persistent blood viral load and an exacerbated inflammatory response[14].

Single-cell omics studies have identified specific transcriptional features in monocytes, natural killer (NK) cells, dendritic cells (DCs) and T cells associated with the severity of COVID-19[13,20–22]. These studies have revealed that severe COVID-19 is marked by a dysregulated myeloid cell compartment[13]. It has also been shown that monocytes from severe COVID-19 patients are characterized by a tolerogenic phenotype with reduced expression of class II major histocompatibility complex (MHC-II) antigens[23] and increased activation of apoptotic pathways[24].

Differentiation and activation of monocytes and other myeloid cells are directly associated with epigenetic mechanisms[25]. The functional plasticity of these cells is also reflected at the epigenetic level and several studies have shown that DNA methylation profiles, among other epigenetic marks, vary in response to inflammatory cytokines, hormones and other factors[26,27], depending on their functionality. Cytosine methylation (5mC) occurs at CpG dinucleotides and is generally associated with transcriptional repression[28], although its relationship with transcription depends on the genomic location of the affected CpG sites[29]. In some cases, DNA methylation changes occur as a result of upstream environmental effects that link cell membrane receptors, signaling pathways and transcription factors (TFs) that can either directly recruit DNA methyltransferases (DNMT) and ten–eleven translocation (TET) enzymes, or indirectly influence their binding to specific genomic sites.

The characterization of the epigenetic and transcriptomic reprogramming in monocytes, given their central role in inflammatory responses, is essential if we are to understand the specific dysregulated pathways involved in severe forms of COVID-19. In this study, we obtained the DNA methylation profiles of peripheral blood monocytes of severe COVID-19 patients and studied their relationship with transcriptomic changes, obtained by generating droplet-based single-cell RNA sequencing (scRNA-seq) data from peripheral blood.

## Methods

### Human Samples

Our study included a selection of 58 severe COVID-19 patients from the Intensive Care Unit (ICU) of Vall d’Hebron University Hospital (Barcelona) recruited during the second wave of infection in Spain (October to November 2020). Peripheral blood samples were taken at different times following admission of the patient to the ICU, as specified in Additional file 1. Table S1 (Days in ICU). 94% of the patients required intubation and all enrolled cases were confirmed to be infected with SARS-CoV-2 by the use of real-time RT-PCR at the time of collection. For all enrolled patients, the date of enrollment, clinical classification or treatment were obtained from the clinical records. From all these patients, 48 of the 58 patients were selected for DNA methylation analysis (Additional file 1. Table S1) and PBMCs from 10 of the 58 patients were used for droplet-based scRNA-seq analysis (Additional file 2. Table S2). The control population for the DNA methylation analysis comprised 11 healthy donors (HDs) recruited at the Blood Bank of Vall d’Hebron University Hospital. Table 1 summarizes the characteristics and clinical data from patients included in the DNA methylation analysis. We included an additional group of 14 patients from the same hospital for DNA methylation and expression validation, including 9 severe COVID-19 patients and 5 mild COVID-19 patients, together with an additional group of 6 HDs. The validation cohort was collected during February 2022, applying the same selection criteria as for the discovery cohort. For the validation cohort, we only included non-vaccinated patients, to match the vaccination status with that of the patients collected in the initial phase of the study. Clinical information corresponding to the new cohort is also included in Additional file 1. Table S1 (validation cohort). This study was approved by the Clinical Research Ethics Committees of Hospital Universitari Germans Trias i Pujol (PI-20-129) and Vall d’Hebron University Hospital (PR(AG)282/2020), both of which adhered to the principles set out in the WMA Declaration of Helsinki. Informed consent was obtained from all patients before their inclusion.

**Table 1.**
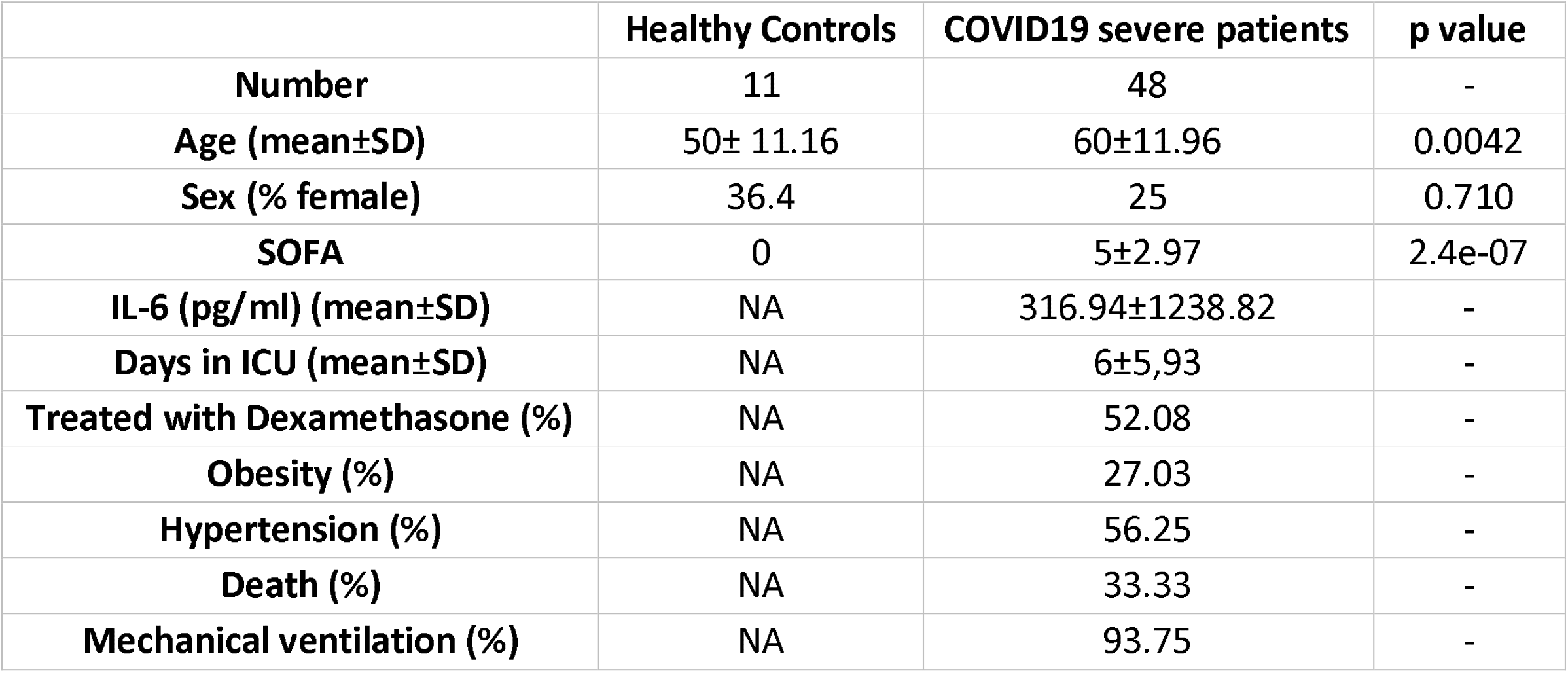
Summary of patient cohort for DNA methylation analysis

|  | Healthy Controls | COVID19 severe patients | p value |
| --- | --- | --- | --- |
| **Number** | 11 | 48 | - |
| **Age (mean±SD)** | 50± 11.16 | 60±11.96 | 0.0042 |
| **Sex (% female)** | 36.4 | 25 | 0.710 |
| **SOFA** | 0 | 5±2.97 | 2.4e-07 |
| **IL-6 (pg/ml) (mean±SD)** | NA | 316.94±1238.82 | - |
| **Days in ICU (mean±SD)** | NA | 6±5,93 | - |
| **Treated with Dexamethasone (%)** | NA | 52.08 | - |
| **Obesity (%)** | NA | 27.03 | - |
| **Hypertension (%)** | NA | 56.25 | - |
| **Death (%)** | NA | 33.33 | - |
| **Mechanical ventilation (%)** | NA | 93.75 | - |

### Monocyte purification and DNA isolation

PBMCs were obtained from peripheral blood by Ficoll gradient using Lymphocyte Isolation Solution (Rafer, Zaragoza, Spain) from 48 of the severe COVID-19 patients and 11 HDs. Once PBMCs were isolated, all samples were stored at -150°C in 10% DMSO in fetal bovine serum (FBS) until monocyte purification. The monocyte population was isolated by flow cytometry (FacsAria Fusion, BD, Beckton Dickinson, San Jose, CA, USA). PBMCs were stained with CD14-APC-Vio770 (Miltenyi Biotec) and CD15-FITC (Miltenyi Biotec) in staining buffer (MACS) for 20 min. A gating strategy was employed to eliminate cell debris, doublets and DAPI+ cells. CD14 and CD15 antibodies were used to isolate CD14+CD15-. Purified cells were pelleted and stored at -80ºC.

After monocyte isolation, DNA was isolated using the AllPrep DNA/RNA/miRNA Universal Kit (Qiagen) following the manufacturer’s instructions.

### DNA methylation profiling

Bisulfite (BS) conversion was performed using EZ-96 DNA Methylation™ Kit (Zymo Research, CA, USA) according to the manufacturer’s instructions. Five hundred nanograms of BS-converted DNA were hybridized on Infinium Methylation EPIC BeadChip arrays (Illumina, Inc., San Diego, CA, USA). These were used to analyze DNA methylation. They enable > 850,000 methylation sites per sample to be assessed at single-nucleotide resolution, which corresponds to 99% of the reference sequence (RefSeq) genes.

Each methylation data point was obtained from a combination of the Cy3 and Cy5 fluorescent intensities from the methylated and unmethylated alleles. Background intensity computed from a set of negative controls was subtracted from each data point. For representation and further analysis, we used *beta* (*b*) and *M* values. *Beta* is the ratio of methylated probe intensity to overall intensity (the sum of the methylated and unmethylated probe intensities). *M* is calculated as the log_2_ ratio of the intensities of the methylated and unmethylated probes. For statistical purposes, the use of *M* is more appropriate since *b*-values are severely heteroscedastic for highly methylated and unmethylated CpG sites. Raw DNA methylation data are available at GEO, with accession number GSE188573.

### Quality control, data normalization and statistical analysis of DMPs

Quality control and analysis of EPIC arrays were performed using ShinyÉPICo[30], a graphical pipeline that uses *minfi* (v1.36)[31] for normalization, and *limma* (v3.46)[32] for analyzing differentially methylated positions. ShinyÉPICo is available as an R package at the Bioconductor (http://bioconductor.org/packages/shinyepico/) and GitHub (https://github.com/omorante/shinyepico) sites. We used the BS conversion control probes test included in ShinyÉPICo to determine whether the conversion rate was above the quality threshold of 2 established by Illumina. The threshold was calculated from the information of the BS conversion control probes of the EPIC arrays. When the BS conversion reaction is successful, control probes display strong signal in the red channel, whereas if the sample has unconverted DNA, control probes have a strong signal in the green channel. The red/green ratio for each control position was calculated for each sample.

CpH and SNP loci were removed by the Noob method, followed by quantile normalization. Sex chromosomes (X and Y) were also excluded from the analysis to avoid discordant information among samples. Even when data were generated in a single batch and randomized, we applied the batch effect correction. Sex and age of the donors were included as covariates, to minimize confounding effects due to differences between the median age of the patient and control cohorts, and the Trend and Robust options were implemented in the eBayes moderated *t*-test analysis. To compare healthy donors with the entire severe COVID-19 patient cohort, we identified differentially methylated CpG sites by using *t*-tests and a method with defined empirical array weights, included in the limma package[32], and selecting CpGs with a false discovery rate (FDR) of < 0.05 and a Δβ of > 0.15. To test the effects of potential changes in monocyte subset proportions, we also included this information as a covariate, and performed the same analysis as above, but including only those samples for which such information was available.

We used the iEVORA package (v1.9.1) [33] to identify differentially variable positions (DVPs). This algorithm identifies differences in variance using Bartlett’s test (FDR < 0.001), followed by the comparison of means using *t*-test (p < 0.05) to regularize the variability test, which is overly sensitive to single outliers. For the analysis in Figure 2, we calculated Spearman’s correlation coefficient (rho) to measure the association of two variables and thereby identify CpG sites in which DNA methylation was correlated with SOFA in patients with severe COVID-19. We selected the CpG sites for which Spearman’s rho was greater than 0.4 and had an associated value of p < 0.01. Principal component analysis (PCA) of *b*-values from ShinyÉPICo was used to determine the correlations of PCs with clinical variables such as dexamethasone treatment, obesity, hypertension, etc. Pearson correlation coefficients between numerical variables and PCs were calculated. Categorical variables were entered in a linear model together with the PCs, which were considered as a function of the variable.

### Gene ontology, transcription factor enrichment, and chromatin state discovery and characterization

The GREAT (v3.0.0) online tool (http://great.stanford.edu/public/html) was used for gene ontology (GO) analysis, in which genomic regions were annotated by applying adapted basal and extension settings (5 kb upstream, 5 kb downstream, 1000 kb plus distal). GRCh37 (UCSC hg19, Feb. 2009) was used as the alignment genome reference. Annotated CpGs in the EPIC array were used as background. GO terms were considered significant for a > 2-fold change and an FDR < 0.05. Enrichment is represented as -log_2_(FDR). GO categories with p < 0.05 were considered significantly enriched. GO analysis of differentially expressed genes (DEGs) was carried out using the online Enricher gene ontology analysis tool (https://maayanlab.cloud/Enrichr/). GO categories with a > 2-fold change and an FDR < 0.05 were considered significantly enriched.

We used the *findMotifsGenome*.*pt* tool from the motif discovery HOMER software (v4.10.3) to analyze motif enrichment[34]. A flanking window of ± 250 bp from each CpG was applied, and CpGs annotated in the EPIC array were used as background. To determine the location relative to a CpG island (CGI), we used ‘hg19_cpgs’ annotation in the *annotatr* (v1.8) R package. The statistical test used for the enrichment in these analyses was Fisher’s exact test. Chromatin functional state enrichment of DMPs was measured using public PBMC data from the Roadmap Epigenomics Project (http://www.roadmapepigenomics.org/) generated with ChromHMM (v1.23)[35]. Enrichments were calculated with a Fisher’s exact test using array annotation as background regions. Only significantly enriched states are shown.

### Heatmaps and PCA plots

Heatmaps of DMPs were generated with functions available in the *ComplexHeatmap* (v2.11.1) and *gplots* (v3.1.3) R packages. We used PCA for the low-dimensional analyses. PCA projection matrices were calculated with R’s prcomp function and visual representations of PCs were plotted with the *ggfortify* package (v4.1.4).

### Whole-genome bisulfite sequencing (WGBS) analysis

DNA methylation values of *Ensembl Regulatory Build* regions of progenitor cells such as hematopoietic stem cell (HSC), multipotent progenitor (MPP), common myeloid progenitor (CMP), granulocyte macrophage progenitor (GMP), and control monocytes were extracted from public whole-genome bisulfite sequencing (WGBS) (GSE87197)[36]. Using GenomicRanges (v1.42.0) and based on genomic location, the overlap of the hypermethylated DMPs observed in COVID-19 compared with HD was determined with the *Ensembl Regulatory Regions* from the hematopoietic precursors and monocytes. For this analysis, all DNA methylation data were annotated with respect to the GRCh38 human genome reference.

### Single-cell capture

PBMCs from 10 ICU patients were used to generate single-cell gel beads-in-emulsion (GEMs) (Additional file 2. Table S2). Cells were then washed three times and counted. For samples with low viability (< 90%), we performed Ficoll separation in an Eppendorf tube to eliminate dead cells and increase cell viability. For samples with greater than 90% viability, we filtered using a Flowmi strainer and counted the cells before loading into 10X chromium to generate single-cell GEMs, following the manufacturer’s instructions. We loaded 50,000 cells per pool, including a total of 4 patients per pool. Datasets from patients and HDs are available as h5ad files (https://www.COVID-19cellatlas.org/index.patient.html (Additional file 2. Table S2). In parallel, genomic DNA was isolated from the same 10 PBMCs for genotyping and subsequent donor deconvolution (as described in[37]) using a Maxwell® 16 Blood DNA Purification Kit from Promega following the manufacturer’s instructions.

### scRNA-seq cell type identification and annotation

Single-cell transcriptome data from COVID-19 patients were quantified and aligned using Cell Ranger (v3.1) with the GRCh38 genome concatenated to SARS-Cov-2 genome as a reference. Thereafter, cells from pooled samples were deconvolved and demultiplexed using *Souporcell* (v3.0)[38], yielding a genotype variant that allows donor identity to be matched across different samples. This additionally enabled the removal of doublet cells that could not be explained by any single genotype. *Scrublet* (v0.2.3) [39] was subsequently employed to further filter out other doublets based on computed doublet scores. Specifically, Student’s *t*-test (p < 0.01) after Bonferroni correction was used within fine-grained sub-clustering of each initial cluster produced by the Leiden algorithm. Data were not denoised because no significant contamination or ambient RNA was present. Previously described scRNA-seq datasets of healthy individuals (HD)[40] were then integrated for comparison using single-cell variational inference (scVI)[41] with a generative model of 64 latent variables and 500 iterations. More specifically, scVI employs a negative binomial model using raw counts, selecting 5,000 highly variable genes to produce the latent variables. Defined cell-cycle phase-specific genes in the *Seurat* package (v4.1.0)[42] were excluded from these to reduce the dependence of clustering on cell-cycle effects. Data were subsequently analyzed using *Scanpy* (v1.9.1)[43] following the recommended standard practices. For quality control, genes expressed in fewer than three cells, and cells with fewer than 200 genes or more than 20% mitochondrial gene content, were removed prior to downstream analysis. Data were normalized (scanpy.pp.normalize_per_cell, scaling factor = 10000) and log_2_-transformed (scanpy.pp.log1p). For gene expression visualization (e.g., heat maps), data were further scaled (scanpy.pp.scale, maximum value = 10).

### Cell type clustering and annotation

The resulting latent representation from the integrated datasets was used to compute the neighborhood graph (scanpy.pp.neighbors), then the Louvain clustering algorithm (scanpy.tl.louvain, resolution = 3) and Uniform Manifold Approximation and Projection (UMAP) visualization (scanpy.tl.umap) were employed. Cell type annotations were manually refined using literature-driven, cell-specific marker genes. Identified residual RBCs from incomplete PBMC isolation were excluded before further analysis, as recommended[44].

### Differential gene expression and transcription factor-enrichment analysis

Differential gene expression between COVID-19 patients and healthy individuals (FDR < 0.05) was analyzed using the *limma* package[45]. To predict transcription factor (TF) involvement in transcriptomic changes we used DoRothEA (Discriminant Regulon Expression Analysis) v2 tool[46]. Regulons with a confidence score of A–C were analyzed, and cases with p < 0.05 and a normalized enrichment score (NES) of ± 2 were considered significantly enriched.

### Cell–cell communication

Based on the differential expression analysis, CellPhoneDB[47] v3 (www.CellPhoneDB.org) was used to infer changes in ligand/receptor interactions between the identified cell types in COVID-19 versus HD. Specifically, instead of random shuffling, as used in the previously described statistical method, differentially expressed genes (FDR < 0.05) were used to select interactions that were significantly enriched in either severe COVID-19 patients or healthy individuals relative to the other group. An interaction was considered enriched if at least one of the two partners (ligand or receptor) was differentially expressed, and if both partners were expressed by at least 10% of the interacting cells.

### Bisulfite pyrosequencing

EZ DNA Methylation-Gold kit (Zymo Research), was used to BS-converted 500 ng of genomic DNA following the manufacturer’s instructions. BS-treated DNA was PCR-amplified using IMMOLASE DNA polymerase kit (Bioline). Primers used for the PCR were designed with PyroMark Assay Design 2.0 software (Qiagen) (Additional file 3. Table S3). PCR amplicons were pyrosequenced with the PyroMark Q24 system and analyzed with PyroMark Q48 Autoprep (Qiagen).

### Real-time quantitative Polymerase Chain Reaction (RT-qPCR)

The Transcriptor First Strand cDNA Synthesis Kit (Roche) was used to convert 250 ng of total RNA to cDNA following the manufacturer’s instructions. RT-qPCR primers were designed with Primer3 software[48] (Additional file 3. Table S3). RT-qPCR reactions were prepared with LightCycler 480 SYBR Green I Master (Roche) according to the manufacturer’s instructions and analyzed with a LightCycler 480 instrument (Roche).

### Flow cytometry

To study the surface cell markers on monocytes (CD14+), PBMCs from the 10 patients used for single cell analysis and 10 HDs were defrosted and washed once with PBS. After blocking for non-specific binding with Fc block (BD Pharmingen) for 5 min on ice, cells were incubated for 20 min on ice using staining buffer (PBS with 4% fetal bovine serum and 0.4% EDTA). Antibodies used included: CD14-FitC (Miltenyi Biotec), CD85-PEvio770 (Miltenyi Biotec), CD172a-APC (Miltenyi Biotec), CD97-PEvio770 (Miltenyi Biotec), CD31-PE (Miltenyi Biotec), CD366-PEvio615 (Miltenyi Biotec), CD62L-APC (Miltenyi Biotec), CD58-PE (Miltenyi Biotec), CD191-PEvio770 (Miltenyi Biotec), CD52-PEvio615 (Miltenyi Biotec), CD48-APC (Miltenyi Biotec). Cells were analyzed in a BD FACSCanto-II flow cytometer.

### Statistical analysis

All statistical analyses were done with R v4.0.2. Box, bar, violin, bubble and line plots were generated using functions from the *ggplot2* (v3.3.6) and *ggpubr* (v4.0) packages. Mean normalized DNA methylation values were compared using two-tailed test. Multivariate frequency distributions were calculated using Fisher’s exact test. The levels of significance are indicated as: * p < 0.05, ** p < 0.01, *** p < 0.001 and **** p < 0.0001.

## Results

### DNA methylome remodeling in peripheral blood monocytes of severe COVID-19 patients

To directly inspect epigenetic alterations in peripheral blood monocytes in severe COVID-19, we isolated CD14+CD15-cells from 59 blood samples, comprising 48 severe COVID-19 patients and 11 healthy donors (HDs), and performed DNA methylation profiling (Figure 1A, Table 1 and Additional file 1. Table S1). For cell sorting, we first separated live cells from debris, then extracted singlets and isolated CD14+CD15-cells to avoid neutrophil contamination (Figure 1B)[49]. Since we selected CD14+ cells, the purification procedure only included classic (CM) (CD14+CD16-) and intermediate monocytes (IM) (CD14+CD16+), excluding the non-classic monocyte (NCM) (CD14lowCD16+) subpopulation, which in healthy individuals corresponds to around 5% of the total monocyte compartment[50]. Negative selection using CD15 was necessary, as there is a significant increase in the frequency of neutrophils in severe COVID-19 patients, as activated neutrophils are not separated in the Ficoll step[51] (Additional file 4. Figure S1A-S1C). To confirm the purity of our monocytes, we performed FACS analysis and obtained an average purity of 98% (example in Additional file 4 Figure S1D). Studies in various other inflammatory diseases have shown that the proportions of monocytes can shift between the three major subsets, i.e., CM, IM and NCM. For instance, it has been shown that severe COVID-19 patients feature reduced NCM and IM populations[52]. The analysis of monocyte subpopulations in our cohort showed a significant increase in the CM population and a decrease in the NCM population (Additional file 4. Figure S1E-S1F). Since we purified CD14+ monocytes, our study only included CM and IM.

**Figure 1.**
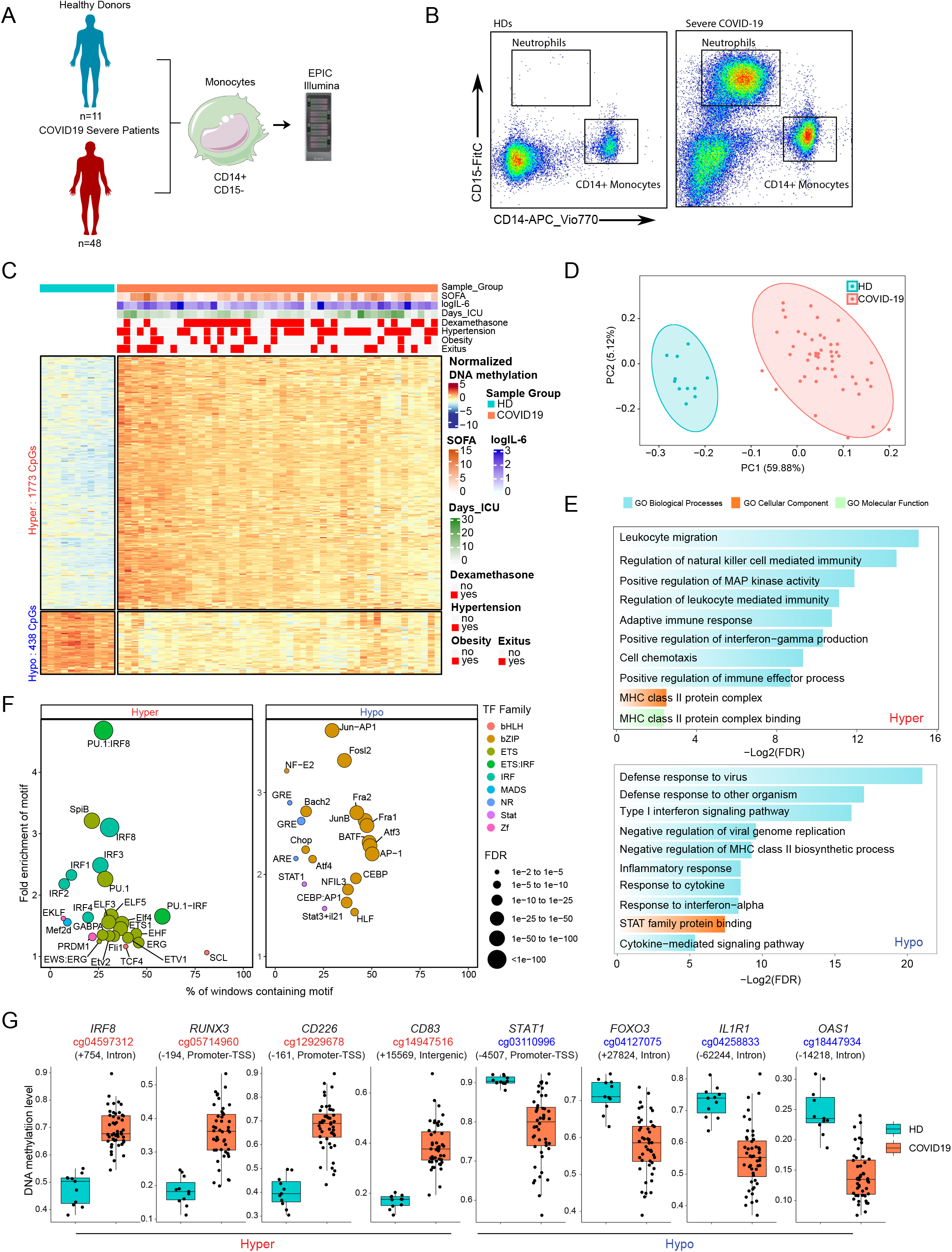
Analysis of DNA methylation in blood monocytes of severe COVID-19 patients. (**A**) Scheme depicting the cohort and workflow for monocyte purification of severe COVID-19 patients and controls and DNA methylation analysis. (**B**) Representative flow cytometry profile, indicating sorting gates used to purify monocytes from HD and COVID-19 patients’ peripheral blood. (**C**) Scaled DNA methylation (z-score) heatmap of differentially methylated positions (DMPs) between HDs (blue bar above) and COVID-19 patients (red bar above). Significant DMPs were obtained by applying a filter of FDR > 0.05 and a differential of beta value (Δß) > 0.15. A scale is shown on the right, in which blue and red indicate lower and higher levels of methylation, respectively. Clinical and treatment data of COVID-19 patients are represented above the heatmap. SOFA, IL-6 level and days in the ICU scales are shown on the right of the panel (**D**) Principal component analysis (PCA) of the DMPs. HDs and severe COVID-19 patients are illustrated as blue and red dots, respectively. (**E**) Gene ontology of hypermethylated and hypomethylated DMPs. Selected significant functional categories (FDR < 0.05) are shown. (**F**) Bubble plot of TF motifs enriched on hypermethylated and hypomethylated DMPs. Bubbles are colored according to their TF family; their size corresponds to the FDR rank. (**G**) Box plot of individual DNA methylation values of CpG from hypermethylated and hypomethylated clusters (*b*-values), with the name of the closest gene and the position relative to the transcription start site.

We performed DNA methylation profiling of isolated monocytes and identified 2211 differentially methylated positions (DMPs) of CpGs in severe COVID-19 patients compared with HDs (FDR < 0.05 and absolute Δß > 0.15). Of these, 1773 were hypermethylated (hypermethylated cluster) and 438 were hypomethylated (hypomethylated cluster) (Figure 1C and Additional file 5. Table S4). PCA of these DMPs showed that the two groups of monocytes (COVID-19 and HD) separated along the first principal component axis (Figure 1D). We obtained similar results when we included monocyte subpopulation proportions as a covariate in the analysis (overlap, p <0.0001) (Additional file 6. Figure S2A). No significant differences (FDR < 0.05) were observed within COVID-19 patients separated by their condition (obesity, hypertension, days admitted to the ICU and exitus/death) or treatment with dexamethasone (Additional file 1. Table S1). None of the abovementioned conditions was significantly correlated with the DNA methylation changes (Additional file 6. Figure S2B). This was also apparent from the PCA showing the overlap of patients with different clinical parameters (Additional file 6. Figure S2C).

The analysis of the genomic functional features of the DMPs in the hypermethylated and hypomethylated clusters (Additional file 6. Figure S2D) using public data from monocytes[35] revealed an enrichment in promoters and enhancers. This is consistent with their proposed roles for DNA methylation in regulatory elements [53].

Gene ontology analysis (GO) of the two DMP clusters revealed several functional categories associated with the immune response to viral infection (Figure 1E). In the hypermethylated cluster, we observed enrichment of categories such as natural killer-mediated immunity, leukocyte migration, adaptive immune response, and positive regulation of interferon gamma production. We also observed hypermethylation in the MHC-II protein complex that was related to antigen presentation. In addition, we found an enrichment of the positive regulation of MAP kinase activity category (Figure 1E, top panel). In the hypomethylated cluster, we observed enrichment of functional categories relevant to viral infection, including defense response to virus and negative regulation of viral genome replication. Importantly, the hypomethylated cluster also featured enrichment of functional categories related to type I interferons (IFN) signaling and MHC class II (Figure 1E, bottom panel).

Transcription factor (TF) binding motif enrichment analysis, in 250-bp windows surrounding DMPs, revealed overrepresentation of TFs of significance to the immune response. The hypermethylated cluster CpGs displayed enrichment of binding motifs of IRFs and ETS TF families, which are linked to IFN changes (Figure 1F, left panel). Motifs of the bZIP TF family like AP-1, Jun, Fosl2, Fra1 and Fra2 were enriched in the hypomethylated cluster. DMPs of the hypomethylated cluster were also enriched in motifs of the signal transducer factor and activator of transcription factor (STAT) members STAT1 and STAT3. We also detected enrichment of the glucocorticoid response element (GRE) in the hypomethylated cluster (Figure 1F, right panel). Given these results, we hypothesized that pharmacological treatment with glucocorticoids (GCs) in severe COVID-19 patients in the intensive care unit (ICU) might influence DNA methylation in monocytes. To test this possibility, we performed *limma* analysis and subsequent binding motif enrichment after separating COVID-19 patients into two groups, with and without GC treatment. Both groups of patients exhibited significant enrichment of GRE motifs in the hypomethylated cluster (Additional file 6. Figure S2E), suggesting that the endogenous production of GCs in severe COVID-19 patients could participate in the hypomethylation through GRE. However, given the size of the cohort we cannot rule out the possibility that pharmacological treatment could also influence DNA methylation changes and therefore remains as a potential confounder factor.

Inspection of the individual genes within or in the vicinity of the DMPs revealed several genes with functions essential to the viral immune response. The list of relevant genes included *IRF8, RUNX3, CD226* and *CD83* in the hypermethylated cluster, and *STAT1, FOXO3, IL1R1* and *OAS1* in the hypomethylated cluster (Figure 1G). We validated these results using bisulfite pyrosequencing in a new cohort of severe COVID-19 patients (Additional file 6. Figure S2F). Interestingly, these changes were also observed in mild COVID-19 patients (Additional file 6. Figure S2F). *IRF8, IL1R1* and *CD83* are associated with the IFN response. *CD226* encodes a glycoprotein related to monocyte, NK and T cell adhesion. This glycoprotein has been shown to be involved in the cytotoxicity of these cells and is known to be altered in COVID-19 patients[13]. *STAT1* is associated with the cytokine response, which, in turn, is related to *IL1R1*. The latter is the receptor of interleukin 1, which participates in the inflammatory response and is strongly expressed in severe COVID-19 patients[14]. *OAS1* is induced by interferons and activates latent RNase, causing viral RNA degradation, which could be related to the identification of the category negative regulation of viral genome replication in the GO analysis.

### Monocytes from severe COVID-19 patients display increased DNA methylation variability

Overall, our DNA methylation analysis showed greater heterogeneity (different variable positions, DVPs) in the profiles from COVID-19 patient monocytes than in those from HDs (Additional file 6. Figure S2G). We then examined the relationship between the DNA methylation profiles and the Sequential Organ Failure Assessment (SOFA) score, which is used in ICUs to calculate organ damage. The score ranges from 0 to 24, with values greater than 6 being associated with a significant increase in the risk of mortality[55]. Using Spearman’s correlation coefficient to assess specific hypermethylated or hypomethylated CpGs with SOFA, we identified 1375 CpG sites whose methylation levels positively correlated with SOFA (increased methylation) (rho < 0.4□and p < 0.01) and 1497 CpG sites with an inverse correlation with SOFA (decreased methylation) (rho <−0.4□and p <0.01) (Figure 2A and Additional file 7. Table S5). The mean normalization DNA methylation profiles of increased and decreased methylation CpG sites were similar in patients with low SOFA (<6) and in healthy controls in an unsupervised representation but differed between the low and high SOFA score groups (Figure 2B). These results suggest that changes in DNA methylation are concomitantly exacerbated for higher SOFA scores, which is associated with bad prognosis. Several CpGs correlating with SOFA were associated with genes, such as *IL17R, SOCS5* and *PCDHA5*, that are involved in T cell-mediated inflammatory responses (Figure 2C). Others, like *FOXG1* and *CDC20B*, are associated with DNA damage. GO analysis revealed that changes in DNA methylation that are concomitant with SOFA show an overrepresentation of terms associated with IFN-γ, production of the molecular mediator involved in inflammatory response, viral gene expression, the B-cell proliferation involved in immune response and Th1 cell cytokine production (Figure 2D).

**Figure 2.**
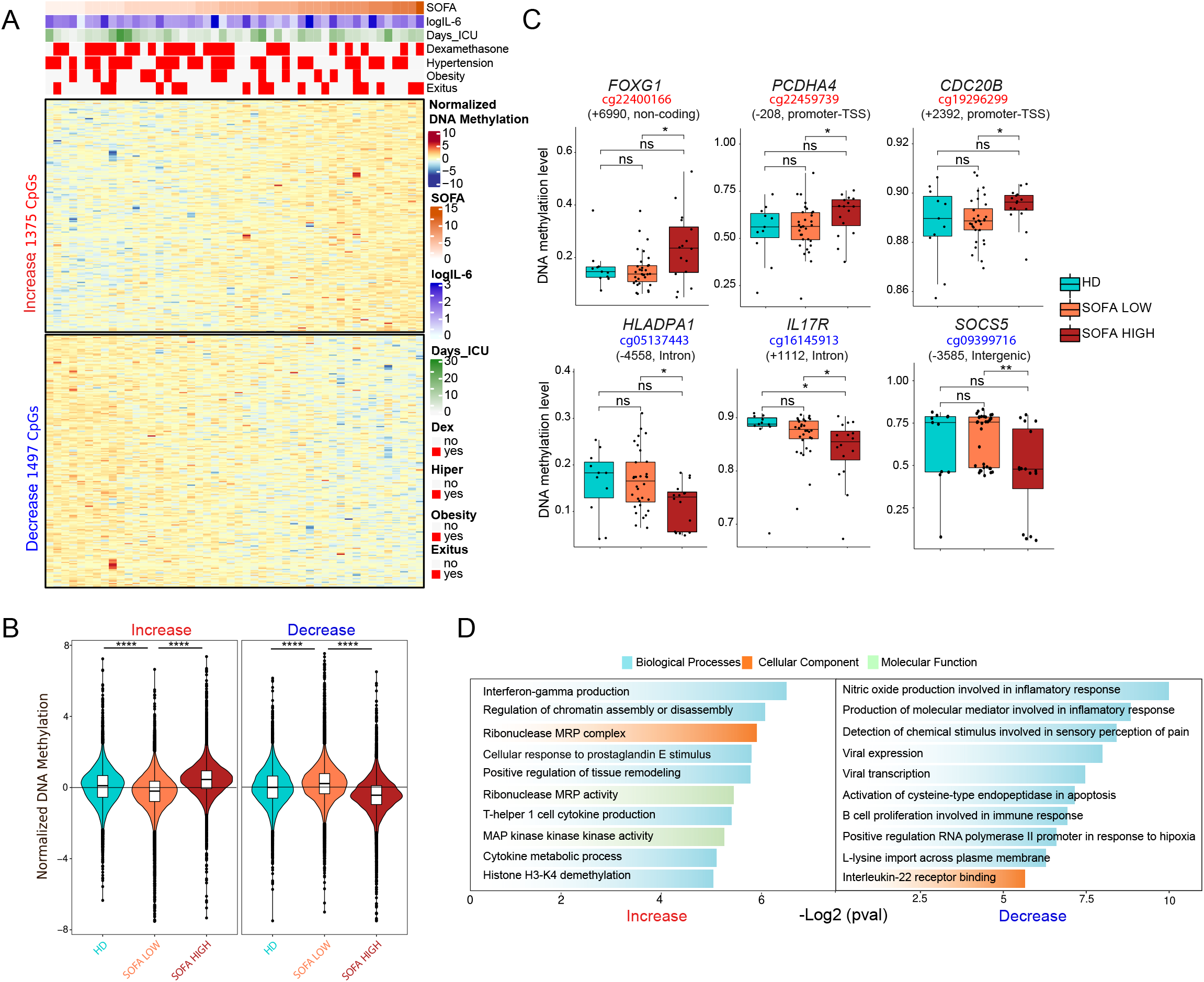
DNA methylation changes in COVID-19 monocytes parallel organ damage. (**A**) Heatmap of severe COVID-19 patients with DNA methylation ordered by SOFA score, including all CpG-containing probes significantly correlated with the SOFA score (Spearman correlation coefficient rho > 0.4, p <□0.01). Clinical and treatment data of COVID-19 patients are shown above the heatmap. SOFA, IL-6 level and days in the ICU scales are shown on the right of the panel (**B**) Normalized methylation values from heatmap showing overall group methylation of HD. Patients with SOFA ≤ 6 are indicated as SOFA LOW; those with SOFA > 6 are indicated as SOFA HIGH. (**C**) DNA methylation levels (*b*-values) of selected individual CpGs (and closest genes) in hypermethylated and hypomethylated sets and their position relative to the transcription start site. Gene ontology (GO) analysis of hypermethylated and hypomethylated DMPs, analyzed with the GREAT online tool, in which CpG annotation in the EPIC array was used as background. Statistical significance: * p < 0.05, ** p < 0.01, *** p < 0.001, **** p < 0.0001.

### DNA methylation alterations in monocytes of severe COVID-19 patients significantly associate with those derived from patients with bacterial sepsis, myeloid differentiation and the influence of inflammatory cytokines

To better characterize the impact of DNA methylation changes in COVID-19, we compared the DMPs from severe COVID-19 patients with those obtained from monocytes derived from patients with bacterial sepsis in a previous study by our team[27], given that severe COVID-19 can be considered a form of sepsis[56]. To this end, we first estimated the DNA methylation values of DMPs corresponding to the sepsis relative to the HD comparison from our previous sepsis study (accession number GSE138074) [27] using the data from the severe COVID-19 methylation dataset. Overall, we found significant enrichment in the hypermethylation and hypomethylation clusters (Figure 3A). We also calculated the odds ratio of the overlap between these two datasets and found a strong enrichment of the hyper-DMPs in COVID-19 relative to those in sepsis (FDR ≤ 2.22·10^−16^) and in the hypo-DMPs (FDR ≤ 2.22·10^−16^) (Figure 3B). We also confirmed an enrichment in introns and depletion in promoters relative to the background when testing the genomic location of the DMPs common to both COVID.19 and sepsis (Figure 3C and Figure S3A). DMPs located in introns are often localized in enhancer regions involved in long-distance regulation[53].

**Figure 3.**
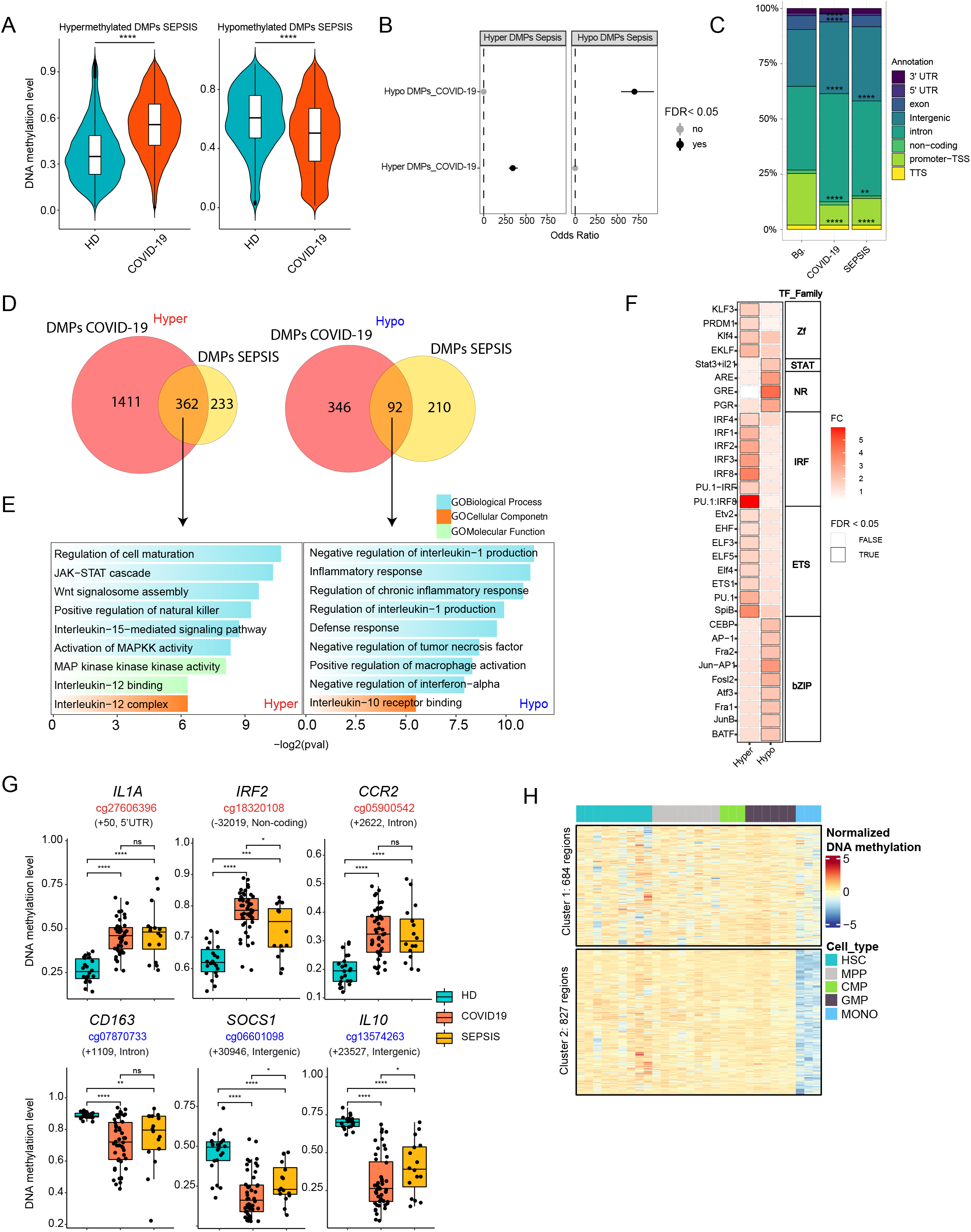
Comparative analysis of DNA methylation in blood monocytes of severe COVID-19 and bacterial sepsis patients. (**A**) Violin plot representing the mean methylation state of the DMPs found in the comparison between HDs and sepsis patients with *b*-values obtained from severe COVID-19 patients. (**B**) Fisher’s exact test showing the odds ratio ± 95% confidence interval of the overlap between DMPs found in monocytes from bacterial sepsis patients and DMPs in monocytes from COVID-19 patients. (**C**) Proportions of the genomic locations (in relation to genes) of DMPs in COVID-19 and sepsis; Bg., background, EPIC probes (**D**) Venn diagram of the overlap of COVID-19 DMPs identified by the comparison of HDs and severe COVID-19 patients with DMPs identified by the comparison between HDs and sepsis patients, separating hypermethylated and hypomethylated DMPs. (**E**) Gene ontology analysis of hypermethylated and hypomethylated overlapping DMPs identified in the previous comparison. Selected significant categories (p < 0.05) are shown. (**F**) TF binding motif analysis of shared hypermethylated and hypomethylated DMPs comparing HDs and COVID-19 patients, and by HDs and sepsis patients. The panel shows the fold change (FC), TF family. Boxes with black outlines indicate TF binding motifs with FDR < 0.05. (**G**) Box-plot showing the DNA methylation values of individual CpGs (together with the name of the closest gene and its position relative to the transcription start site) from the hypermethylated and hypomethylated clusters from both COVID-19 and sepsis. (**H**) Scaled DNA methylation heatmap of regions from the whole-genome bisulfite sequencing (WGBS) data of hematopoietic stem cells (HSCs), multipotent progenitors (MPPs), common myeloid progenitors (CMPs) and granulocyte macrophage progenitors (GMPs) that overlap with the genomic location of the 1772 hypermethylated DMPs identified in the COVID-19 vs. HDs comparison. Statistical significance: * p < 0.05, ** p < 0.01, *** p < 0.001, **** p < 0.0001.

We then determined that the two datasets had 362 hypermethylated and 92 hypomethylated CpGs in common (Figure 3D), corresponding to 51% of the total DMPs of the sepsis patients (Additional file 8. Figure S3B). GO analysis of the shared DMPs revealed significant enrichment in functional terms related to host response, including regulation of NK cells, inflammatory response and leukocyte chemotaxis (Additional file 8. Figure S3C). Shared hypermethylated CpGs were enriched in functional categories related to cell signaling, such as the JAK-STAT and MAPK pathways, that could be involved in the reduction of the inflammatory response and the IL15- and IL12-mediated signaling pathways, which are related to cytokine production and Th1 proliferation (Figure 3E, left panel). Shared hypomethylated CpGs were enriched in functional categories responsible for regulating the inflammatory response, such as negative regulation of IL-1 production and positive regulation of macrophage activation. In concordance with the hypermethylated cluster, we also observed negative regulation of IFNα production (Figure 3E, right panel). It is of note that severe COVID-19-specific DMPs were enriched in functional categories related to virus infection, such as the defense response to virus, and impairment of the antigen-presenting process, which seems to be specific to COVID-19 infection[13,23] (Additional file 8. Figure S3D).

Inspection of TF binding motifs corresponding to the DMPs shared between the two groups, separating the shared hypermethylated and hypomethylated CpG sets revealed IRF family transcription factors like IRF1, IRF2, IFR3 and IRF8 in the shared hypermethylated CpG set, which are well established regulators of the type I IFN system, being common in viral and bacterial infections[57]. We also detected enrichment of the ETS transcription factors that are regulated by MAPK proteins, which were enriched in the GO analysis (Figure 3F). In the shared hypomethylated set, we noted enrichment of STAT3 and TFs from bZIP AP-1, like Jun, and other bZIPs, like CEBP. Interestingly, GRE was also present in the shared hypomethylated cluster (Figure 3F). This suggests the influence of GC in the acquisition of aberrant methylation profiles in COVID-19 and sepsis. Individual genes associated with the COVID-19/sepsis shared hypermethylated and hypomethylated CpG genes include type I IFN-related genes, like *IRF2*, and others, such as *IL1A* and *CCR2*, that are involved in inflammatory processes and monocyte chemotaxis, respectively (Figure 3G). We also identified several genes among the shared hypomethylated set, like *CD163, SOCS1* and *IL10*, that have been associated with the acquisition of tolerogenic properties[58] (Figure 3G).

In both infections, systemic inflammation could be responsible for part of the DNA methylation changes that arise in monocytes. To address this possibility, we examined the DNA methylation levels of the hypomethylated and hypermethylated CpGs of severe COVID-19 and sepsis patients in monocytes isolated from healthy donor PBMCs that had been treated *in vitro* with inflammatory cytokines like IFNα, IFNγ and TNFα[26] (accession number GSE134425). This analysis revealed several significant changes following the trends for both COVID-19 and sepsis (Additional file 8. Figure S3E), suggesting that these inflammatory cytokines, which are elevated in these patients, could influence the monocyte DNA methylomes.

An alternative explanation for the observed changes in severe COVID-19 monocyte methylomes could be that DNA methylation changes reflect alterations during myeloid/monocyte differentiation or the release of immature or aberrant monocytes. This has been described in severe COVID-19 cases[13,59–62]. It is worth noting that immature cells are also released from the bone marrow in sepsis[63]. To test this hypothesis, we used public whole-genome bisulfite sequencing (WGBS) data (GSE87197) of progenitor cells including HSC, MPP, CMP and GMP cells and monocytes as references. We compared the 1773 hypermethylated CpGs based on their genomic location and obtained 1511 unique Ensembl Regions, which grouped in two clusters. Cluster 1 showed low-level demethylation in monocytes compared with all hematopoietic precursor cell types, whereas cluster 2 showed clear demethylation in monocytes (Figure 3H). These results are compatible with the possibility that a proportion of the DMPs in severe COVID-19 result from aberrant myeloid differentiation or the release of immature monocytes, which display higher methylation levels, and are not demethylated to the extent they are during normal differentiation.

### Aberrant DNA methylation is associated with changes in gene expression of COVID-19 patient monocytes

To study the relationship between the DNA methylation changes and aberrant gene expression of monocytes derived from severe COVID-19 patients, we obtained single-cell (sc) RNAseq data of peripheral blood mononuclear cells (PBMCs) from 10 additional severe COVID-19 patients from the same hospital and compared them with those of 10 HDs from a public dataset[40] (Additional file 2. Table S2 and Additional file 9. Figure S4A-S4B). This analysis enabled us to identify 24 cell populations based on specific markers (Figure 4A and Additional file 9. Figure S4C-S4D), and thereby not only to determine the alterations in gene expression in monocytes, but also to inspect alterations in additional immune cell subsets. Strikingly, the monocyte fraction comprised solely CD14+ cells (CD14 mono: *CD14*) (Figure 4B).

**Figure 4.**
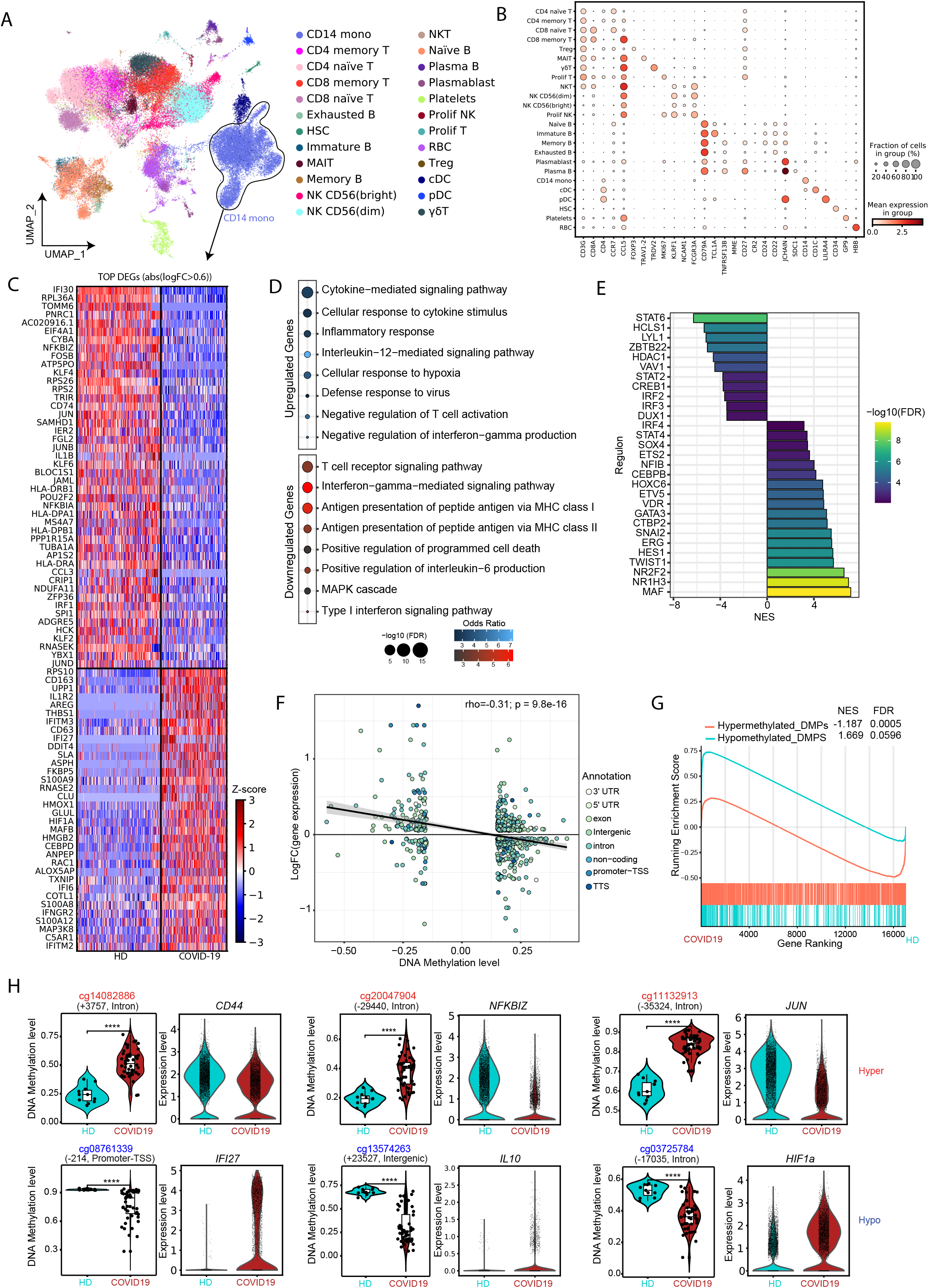
Correlation between DNA methylation and gene expression. (**A**) UMAP visualization showing the immune cell populations identified from Louvain clustering and cell-specific marker gene expression. (**B**) Dot plot representing the expression of selected marker genes identified in the cell population. The scale represents the mean gene expression level in the cell subset and the circle size represents the percentage of cells in the subset of expressing cells. (**C**) Heatmap representing differentially expressed genes (DEGs) with a log_2_(FC) > 0.6, above, and log_2_(FC) < - 0.6, below. Genes overexpressed and downregulated in COVID-19 patients in relation to HDs are depicted in red and blue, respectively. (**D**) Gene ontology (GO) over-representation of GO Biological Process categories comprising the upregulated and downregulated DEGs. The odds ratios for each group and the -log_2_(FC) are shown. Selected significant categories (FDR < 0.05) are shown. (**E**) Discriminant regulon expression analysis (DoRothEA) of COVID-19 severe patients compared with HDs. Normalized enrichment score (NES) and log_2_(FC) of transcription factor expression are depicted. (**F**) Correlation of average DNA methylation levels of DMPs with average gene expression of DEGs in the HDs *vs*. COVID-19 severe patients. Log_2_(FC) of expression is plotted on the y-axis, higher numbers representing a higher level of expression in COVID-19 and lower numbers a higher level of expression in HDs. DNA methylation is depicted on the x-axis as Δβ, lower numbers representing a lower level of methylation in COVID-19 monocytes, and higher numbers a lower level of methylation in HDs. Points are colored according to their genomic context. (**G**) Gene set enrichment analysis (GSEA) of HD *vs*. COVID-19, using hypomethylated-associated genes and hypermethylated-associated genes as genesets. The running enrichment score is represented, and the normalized enrichment score (NES) is shown above (FDR < 0.01) (**H**) Representation of individual DNA methylation values of DMPs from the hypermethylated and hypomethylated clusters (*beta* values), the position in respect to the transcription start site, and the relative expression of the closely related DEGs. Statistical significance: * p < 0.05, ** p < 0.01, *** p < 0.0001, **** p < 0.00001.

In the CD14+ monocyte cluster, we identified 10440 differentially expressed genes (DEGs) between COVID-19 patients and HDs (Additional file 10. Table S6). The top DEGs (based on the fold change (FC)), included pro-inflammatory molecules *(IL1B, CCL3*), surface markers (*CD163, CD63, AREG, CD74, S100A12, S100A12, S100A8, S100A9*) and transcription factors (*JUN, MAFB, NF-KB*) (Figure 4C). We observed upregulation of monocyte-derived cell markers like *S100A12, S100A8* and *S100A9. S100A8* is already known to contribute to the cytokine storm in severe COVID-19[40,64]. Proinflammatory genes like *IL1B* of *IRF1* were downregulated; as well as *HLA* genes, in agreement with previous studies, suggesting decreased antigen presentation in severe COVID-19 patients. Finally, we observed downregulation of the NF-κB inhibitor zeta-encoding gene *NFKBIZ*, consistent with activation of this proinflammatory pathway[65]. Since type I IFNs are essential for antiviral immunity, and the DNA methylation analysis had indicated the potential occurrence of epigenetic alterations in IFN-stimulated genes (ISGs), we checked the levels of genes regulated by type I IFNs and found downregulation of several ISGs, such as *STAT1, BST2, PTPN6* and *IRF1* (Additional file 11. Figure S5A). In addition, given that some of the observed DNA methylation changes were associated with genes involved in antigen presentation, we inspected HLA genes in our expression data and found this gene set to be significantly downregulated, consistent with dysfunction in antigen processing and presentation (Additional file 11. Figure S5B).

GO analysis of both DEG sets revealed enrichment in functional terms coincident with those from DNA methylation analysis. We observed functional categories such as cytokine-mediated signaling, IL-12-mediated signaling, negative regulation of T-cell activation, negative regulation of IFNγ production and defense response to the virus in the upregulated cluster genes (Figure 4D). Conversely, functional categories such as antigen processing and presentation by MHC-I and MHC-II and IFNγ-mediated signaling were enriched among the downregulated gene set (Figure 4D). We then studied TFs potentially involved in the transcriptomic changes observed in COVID-19 monocytes, using Discriminant Regulon Expression Analysis (DoRothEA) analysis, and found that MAF family members, GATA3, STAT4 and IRF4 were associated with upregulated genes in severe COVID-19 (Figure 4E). Conversely, STAT6, STAT2, IRF2, IRF3 and LYL1 were associated with downregulated genes (Figure 4E). TF enrichment of upregulated and downregulated genes was also consistent with the results from DNA methylation analysis, in which binding motifs for several of these TFs were overrepresented among the regions neighboring the DMPs.

We determined the significance of a negative correlation between DMPs and the expression levels of their closest genes (rho = -0.31; *p* = 9.8e-16) (Figure 4F). To study the relationship between DNA methylation and expression changes further, we performed Gene Set Enrichment Analysis (GSEA) of the genes associated with hypermethylated and hypomethylated CpG clusters. Genes associated with hypermethylated CpGs were generally downregulated (NES = 1,669; FDR = 0.0005), whereas those associated with hypomethylated CpGs were upregulated (NES = -1,187; FDR = 0.0596) in COVID-19 patients (Figure 4G). GO analysis of genes with an inverse relationship between methylation and expression levels showed enrichment of functional categories like negative regulation of T cells, IFNα and antigen presentation (Additional file 11. Figure S5C-S5D). This analysis reinforced the relationship between DNA methylation changes and expression changes related to the acquisition of a more tolerogenic phenotype in monocytes in COVID-19 patients. Some examples include *IL10*, a tolerogenic cytokine whose expression is increased in COVID-19, and *NFKBIz*, whose level of expression is decreased (Figure 3H). We validated these results using bisulfite pyrosequencing and qRT-PCR with a new cohort of severe COVID-19 patients (Additional file 11. Figure S5E-F). The analysis also included mild COVID-19 that showed partial or total DNA methylation changes to the extent seen in severe COVID-19 cases (Additional file 11. Figure S5E-F).

### Potential relationship between transcriptional and epigenetic reprogramming and altered immune cell–cell communication

Given the overrepresentation of genes associated with cytokine activity, MHC class II-mediated antigen presentation among the observed DNA methylation and gene expression alterations in severe COVID-19, we explored the potential correlation of these changes in monocytes with their pattern of communication with other immune cell types. To systematically analyze the effect of cell– cell communication on monocytes, we used CellPhoneDB (www.cellphonedb.org), a repository of ligands, receptors and their interactions integrated within a statistical framework that predicts enriched cellular interactions between two cell types using scRNA-seq datasets. This allowed us to infer potentially altered interactions between monocytes and other immune cell subsets in severe COVID-19. In particular, we inspected cell–cell communication alterations between CD14+ and CD4+ memory, CD4+ naïve, CD8+ memory and CD8+ naïve T cells; B cell subsets including memory, naïve and plasma B cells; natural killer cells (NK CD56^dim^: NK CD56^bright^) (Figure 5A-B). Our analysis revealed 4483 ligand/receptor pairs, in which the expression levels of ligands and receptors of CD14+ and/or interacting partners in the aforementioned cell types were significantly different between severe COVID-19 patients and HDs, suggesting changes in the interaction of the corresponding immune cells (Additional file 12. Table S7). The aberrant levels of the proteins encoded by these genes in monocytes were validated by flow cytometry (Figure S5G), supporting a potential impact on cell-cell communication.

**Figure 5.**
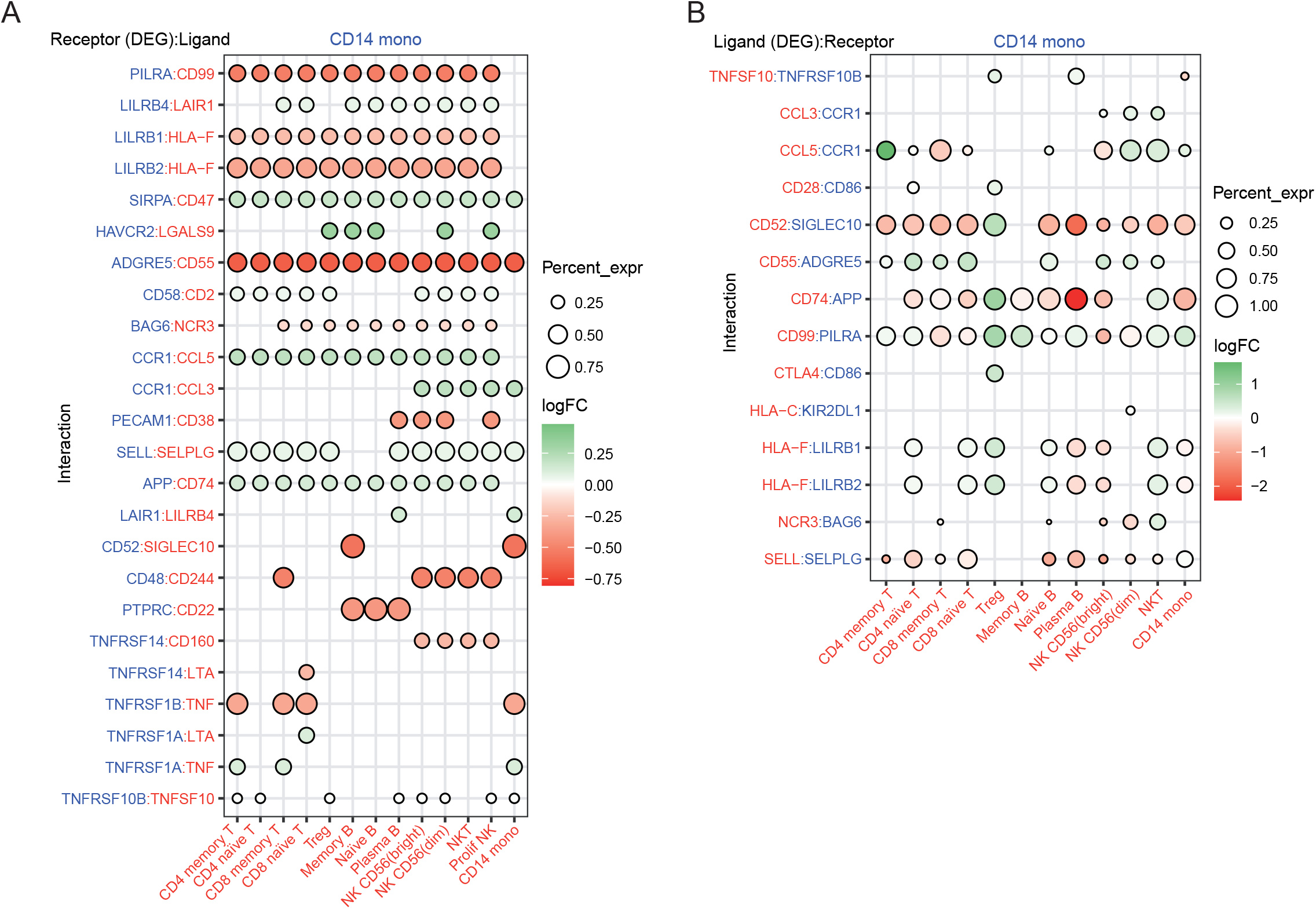
Cell–cell communication analysis. Dot plot of selected receptor/ligand pair (**A**) and ligand/receptor (**B**) interactions between CD14+ monocytes and other cell components in the COVID-19 patient group. Gene expression is indicated as log_2_(FC) for differentially expressed genes (FDR < 0.05), which, in both cases (A and B), are the molecules presented on the left. The percentage expression of the differentially expressed genes in each cell type is indicated by the circle size. Molecules shown in blue are those expressed in CD14+ monocytes. Molecules expressed in the immune cell partner are shown in red.

Figure 5A illustrates the significant ligand-receptor interactions that may be affected when the expression of receptor in monocytes is altered, revealing their potential impact on other cell types. In general, there was a high frequency of interactions involving different types of NK cells, consistent with the terms observed in the GO analysis performed with DMPs (Figure 1E). *PILRA, LILRB1, LILRB2* and *PECAM1* (CD31), the products of which are involved in the inhibition of immune response, were downregulated. Their corresponding ligands, CD99, HLA-F and CD38, were expressed in all the analyzed cell types, except for CD38, which is only expressed in NK and plasma B cells. Additionally, the gene encoding for receptor LAIR1, which inhibits IL-2 expression, was upregulated in monocytes[66], which might influence the interaction with cells expressing its corresponding ligand, i.e., plasma B cells and monocytes. Our analysis also revealed changes in the expression of TNF receptor genes (*TNFRSF14, TNFRSF1B, TNFRSF1A*) in monocytes, which could affect the interaction with T cells through the ligands encoded by *TNF* and *LTA*. This is compatible with the possibility that TNF-associated DNA methylation alterations in monocytes could arise from altered interactions with T cells through these ligand-receptor pairs. We also noted downregulation of the receptor *TNFRSF14*, which interacts with CD160 in NK cells. Some studies have argued that CD160 is essential for NK-mediated IFN-γ production[67], a conclusion that is consistent with the results obtained in our gene ontology analysis of the DNA methylation data. *ADGRE5* (CD97) was downregulated in monocytes. This receptor interacts with CD55, which is expressed in all the analyzed cell types. This interaction is involved in leukocyte migration[68]. The potential alteration of this interaction could be linked to the observed hypomethylation of CpGs close to genes related to leukocyte migration (Figure 1E, top).

We also examined DEGs corresponding to ligands expressed in all immune cell types, whose corresponding receptors are expressed in monocytes, to identify potential cell–cell communication alterations that might affect monocytes (Figure 5B). In general, we detected upregulation of ligands in regulatory T cells (Treg) and downregulation of ligands in plasma B cells. We also observed increased levels of *CCL5* and *CCL3*, expressed in NK cells, that interact with the CD191 receptor (*CCR1*), and whose inhibition potentially suppresses immune hyperactivation in critical COVID-19 patients[69]. In the context of antigen presentation, there was upregulation of HLA-F from Treg and NKT, which interacts with *LILRB1* in monocytes. Recent studies have associated *LILRB1* with the development of tolerance[70]. Our analysis also revealed low levels of CD99, expressed in CD4 memory and naïve T cells, Treg and memory B cells, and the receptor PILRA, which is expressed in monocytes. The opposite occurs with CD8 memory and naïve T cells and NK CD56(bright), which enhances T cell migration[71]. There was a similar trend between CD74 and the receptor APP expressed in monocytes, which is involved in antigen processing and presentation. This could be related to the impaired antigen presentation previously highlighted in our data.

In brief, the potential alteration of cell–cell communication events, through increased or decreased levels of ligands and receptors involving inflammatory cytokines, antigen presentation-related factors and cell activation regulators, in severe COVID-19 patients could affect downstream cell-signaling pathways and TFs, and perhaps influence DNA methylation profiles in monocytes, thereby perpetuating aberrant immune responses.

## Discussion

Our results reveal that peripheral blood monocytes from severe COVID-19 patients display aberrant DNA methylomes and transcriptomes associated with functions related to IFN type I signaling and antigen presentation, among others. The changes are significantly associated with organ damage and with DNA methylation changes occurring in bacterial sepsis. Finally, our analysis suggests that pro-inflammatory cytokines, the release of immature or aberrant monocytes, and specific dysregulated immune cell–cell communication events may be responsible for some epigenetic changes.

To date, there have been very few DNA methylation studies addressing the involvement of COVID-19 DNA methylation in regulating the Angiotensin-converting Enzyme 2 (ACE2) type I membrane receptor gene[72], which is present in arterial, lung type II alveolar cells, where it acts as a SARS-CoV-2 receptor. There is a suggestion that the host epigenome may represent a risk factor for COVID-19 infection. Very few studies have reported alterations in DNA methylation in relation to immune responses[73–75]. Our study aimed to explore the involvement of DNA methylation in relation to a severe COVID-19 outcome in the myeloid compartment, which is directly related to systemic inflammation. We specifically studied monocytes because it is the cell type that undergoes the most dramatic transcriptomic reprogramming during COVID-19 infection[13,21,23,76]. In this regard, our study provides the first instance of DNA methylome profiling in a specific immune cell type in COVID-19 patients.

Our data revealed that most DNA methylation changes in monocytes derived from severe COVID-19 patients occurred in genomic sites enriched in PU.1 binding motifs, consistent with earlier studies showing its role as a pioneer TF directly recruiting TET2 and DNMT3b[77]. In our case, most DNA methylation changes occurred in genes related to cytokines, MHC class II proteins and IFN signaling. Similar results about the defective function of MHC-II molecules and activation of apoptosis pathways were obtained in single-cell atlas studies of PBMCs from severe COVID-19 patients[6,21,78,79] and in sepsis[80,81].

We found that DNA methylation changes in severe COVID-19 patients share some features with sepsis, especially those associated with the expression of tolerogenic cytokines like IL-10[82]. The acute phase of these infections suggests a dysregulated inflammatory host response, resulting in an imbalance between pro-inflammatory and anti-inflammatory mediators[14]. Some studies have suggested that viral components induce STAT1 dysfunction and compensatory hyperactivation of STAT3 in SARS-CoV-2-infected cells[83]. We noted the involvement of kinases like JNK, and earlier studies had shown that COVID-19 infection activates the *JNK* and *ERK* pathways that end in the AP-1-dependent gene expression of proinflammatory cytokines[84]. One of the most strongly affected TFs is STAT2, together with STAT6, which could be linked to the aberrant IFN signaling in monocytes in COVID-19[83]. The presence of STAT2 downregulation also suggests a deficiency in the ability to cross-present to CD8+ T cells [85].

We also identified GRE binding sites in association with DNA methylation changes. Generally, the glucocorticoid receptor (GR) is activated when patients are treated with GC. However, we also noted significant GRE enrichment in patients who were not treated with GC, suggesting that endogenous production of GC in COVID-19 patients could regulate GR and affect DNA methylation at its genomic binding sites. GC is also produced endogenously in sepsis patients in whom cytokines like IL-1β, TNFα and IL-6 induce its production from the adrenal cortex using cholesterol as a substrate to reduce inflammatory responses[86,87]. These cytokines were hypomethylated and overexpressed in our dataset, consistent with the results of other studies that have reported increased levels in the serum of COVID-19 patients[88,89]. GRE binding sites are enriched in the DMPs common to COVID-19 and sepsis. GR is a nuclear receptor expressed in most cell types that can trigger the expression of anti-inflammatory genes through direct DNA binding. Furthermore, GRE represses the action of other inflammation-related TFs, including members of the NF-KB and AP-1 families[90,91], which are also known to be downregulated in our cohort. Taken together, our results suggest the existence of a relationship between extracellular factors associated with the cytokine storm occurring in severe COVID-19 and DNA methylation changes. Several studies have shown an increase in the levels of inflammatory cytokines in severe COVID-19, which may contribute to the severity of the disease[92].

However, it is also possible that the DNA methylation changes are partly due to the release of immature or altered monocytes from myelopoiesis, as reported for severe COVID-19[93–96] and sepsis[63]. Release of immature myeloid cells from the bone marrow in severe COVID-19 is reminiscent of emergency myelopoiesis[97]. This is a well-known phenomenon, characterized by the mobilization of immature myeloid cells to restore functional immune cells, and by its contribution to the dysfunction of innate immunity[98]. In fact, a proportion of the hypermethylated CpGs in monocytes from severe COVID-10 patients overlap with regions that become demethylated during myeloid differentiation. This suggests that part of the hypermethylated CpG sites in isolated peripheral blood CD14+ might be associated with aberrantly differentiated monocytes released into the bloodstream in severe COVID-19 patients. However, the small numbers of CD34+ cells in the PBMC fraction of COVID-19 patients and the lack of CD14+ cells in this subset suggest no interference with our results for CD14+CD15-cells, isolated with our method.

The relationship between DNA methylation and gene expression is complex. DNA methylation patterns are cell-type-specific and are established during dynamic differentiation events by site-specific remodeling at regulatory regions[99]. In general, methylation of CpGs located in gene promoters, first exons, and introns is negatively correlated with gene expression[100]. The analysis of our data shows that there is an inverse correlation between the CpG methylation changes and the expression levels of the closest genes. The comparison of the inferred TFs associated with DNA methylation changes and gene expression changes shows common factors like IRF2 and IRF3, which regulate downregulated genes and hypermethylated CpGs. In this context, it is possible that reduced levels of IFN regulatory factor IRF3 or defective IRF7 function reduces the level of IFNα/β gene expression, increasing the sensitivity to viral infection[12,101].

Finally, analysis of cell–cell communication has revealed potential relationships between DNA methylation changes and altered communication of monocytes and other immune cells (e.g., T, plasma B and NK cells). Our data suggest the potential reduction of interactions between monocytes and NK cells through CD160, which mediates the antibody-dependent cell-mediated cytotoxicity that it is essential for IFNγ production[67]. The potentially greater interaction between monocytes and Treg through multiple ligand and receptor pairs is an interesting finding, since Tregs are immunosuppressive cells responsible for maintaining immune homeostasis[102]. In any case, the use of CellPhone DB is useful for inferring cell-cell communications events; however additional validation experiments would be necessary to validate interactions and activation of downstream signaling pathways.

In our study, we could not determine whether the observed DNA methylation alterations in COVID-19 were the cause or the consequence of the changes in gene expression. The analysis of mild COVID-19 cases, in which the DNA methylation and expression level of a few genes showed differences in their similarities with severe COVID-19 cases, suggests that there are cases where expression changes might anticipate DNA methylation changes. In any case, it is reasonable to propose that some DNA methylation changes help perpetuate dysregulated immune responses.

Some limitations of our study include the size of the cohort, and the unequal numbers of individuals administered particular drugs in the different patient groups, which could have affected the COVID-19 data. However, despite these limitations, we found no significant differences among severe COVID-19 patients with respect to the time they were admitted to the ICU or began to receive treatment. This suggests that DNA methylation is quite a general occurrence in the context of COVID-19. Another limitation concerns the cell population analyzed, since the method for monocyte isolation comprises two populations, CM and IM, one of which (CM) is expanded in the patient group. However, the analysis including the monocyte subsets as a covariate indicates that there are no major differences. Finally, in the comparison with DNA methylation of progenitor cells, it is important to note that the DMPs were overlapped with genomic regions, and not single-base data, and further analyses would be required.

Future studies would benefit from having access to a wider cohort in which it is possible to identify significant links between alterations and drug treatments. Incorporating mild and asymptomatic cases would improve our ability to dissect drug- and severity-related specificity in relation to DNA methylation changes. As is the case for other medical conditions, the analysis of DNA methylation changes would be very likely to help predict disease severity, progression and recovery.

## Conclusions

Our study provides unique insights into the epigenetic alterations of monocytes in severe COVID-19. We have shown that peripheral blood monocytes from severe COVID-19 patients undergo changes in their DNA methylomes, in parallel with changes in expression, and that these significantly overlap with those found in patients with sepsis. We have also shown DNA methylation changes are associated with organ dysfunction. Finally, our results suggest a relationship between DNA methylation changes in COVID-19 patients and changes that occur during myeloid differentiation and others that can be induced by pro-inflammatory cytokines. CellPhoneDB analysis also suggests that alterations in immune cell crosstalk can contribute to transcriptional reprogramming in monocytes, which involves dysregulation of interferon-related genes and genes associated with antigen presentation and chemotaxis.

## Data Availability

DNA methylation data for this publication have been deposited in the NCBI Gene Expression Omnibus and are accessible through GEO SuperSeries accession number GSE188573.

## Availability of data and materials

DNA methylation data for this publication have been deposited in the NCBI Gene Expression Omnibus and are accessible through GEO SuperSeries accession number GSE188573. All raw single-cell data used in this study are available as h5ad files (https://www.COVID-19cellatlas.org/index.patient.html).

## Acknowledgements

We are very grateful to the Cytometry Laboratory and the Scientific and Technological Centers, University of Barcelona (CCiT-UB), for their help with the gating strategy and cell sorting.

## Author contributions

G.G.-T. and E.B. conceived and designed the study; G.G.-T., A.G.F.-B. and L.C. prepared and purified the samples; G.G.-T., O.M.-P., C.C.-F. and R.M.-F. performed bioinformatic analyses; A.B. analyzed the single-cell datasets; A.R.-S., M.M.G., R.F. and J.C.R.-R. provided the patient samples and analyzed the clinical data; G.G.-T., J.R.-U., R.V.-T. and E.B. analyzed and interpreted the data; G.G.-T. and E.B. wrote the manuscript; all authors read and approved the final manuscript.

## Ethics approval and consent to participate

This study was approved by the Clinical Research Ethics Committees of Hospital Universitari Germans Trias i Pujol (PI-20-129) and Vall d’Hebron University Hospital (PR(AG)282/2020), which adhered to the principles set out in the WMA Declaration of Helsinki. All samples were managed in compliance with participants’ written informed consent to participate in the study.

## Consent for publication

Written informed consent for publication was provided by the participants.

